# Prevalence and moderators of apathy after traumatic brain injury: a systematic review and meta-analysis

**DOI:** 10.1101/2025.02.28.25323071

**Authors:** Jessica Lynch, Leila Harriet Sarih, Joseph Mole, Grace Revill, Vaughan Bell

## Abstract

**Background:** Apathy is a recognised neuropsychiatric syndrome in individuals with traumatic brain injury (TBI) with far-reaching consequences, including reduced levels of independence, participation in meaningful activities and quality of life. However, previous studies have reported variable prevalence rates and no meta-analysis has synthesised prevalence findings and identified moderators of apathy in clinical populations.

**Methods:** We conducted a pre-registered meta-analysis (PROSPERO CRD42024552306), searching three databases (APA PsycINFO, MEDLINE, and EMBASE) for primary studies assessing apathy in individuals with TBI. 18 studies met inclusion criteria, and data were extracted for meta-analysis to estimate the pooled prevalence of apathy. Subgroup analyses and meta-regressions explored the influence of potential moderating factors including demographic characteristics, injury-related factors, and methods of apathy assessment. The meta-analysis is available online as a computational notebook with an open dataset.

**Results:** The meta-analysis found the prevalence of apathy following TBI to be 37.6% [95% CI 28.5 – 47.2%]. Significant heterogeneity was observed, with prevalence rates ranging from 4% to 87%. Key moderators included cause of injury, TBI severity, sex and population type. Specifically, transport accidents were associated with higher apathy prevalence, while mild TBI, male sex, and veteran status were associated with lower apathy prevalence.

**Conclusions:** Apathy is a prevalent and significant symptom following TBI, affecting over one-third of individuals in the reviewed studies. These findings highlight the need for increased clinical focus on apathy as an important aspect of TBI recovery.

## Introduction

Traumatic brain injury (TBI) is a significant public health concern. It occurs when an external force damages the brain, which can result in a range of physical, cognitive, emotional, and behavioural impairments (Menon et al., 2010). It is the leading cause of disability and mortality among individuals aged 1 to 45, with only 25% of those who have had a serious TBI gaining long-term functional independence (Ahmed et al., 2017). The consequences of TBI extend beyond the individual, often causing significant distress for families and contributing to broader societal and economic challenges (Faul et al., 2010; Rubiano et al., 2015). In 2019, there were 27.16 million new cases of TBI worldwide, with 48.99 million people living with TBI (Guan et al., 2023). In the UK, a report by the Centre for Mental Health estimated the annual cost of TBI to be £15 billion (Parsonage, 2016). Importantly, while some of the symptoms or TBI may be immediately apparent, other, predominantly neuropsychiatric, symptoms, may be less so. It is because of this that TBI is often referred to as the ‘silent epidemic’.

Apathy is a well-recognised neuropsychiatric outcome in individuals with TBI, with significant impact on functioning and recovery (Azouvi et al., 2017; Ciurli et al., 2011; Worthington & Wood, 2018). Marin (1991) first defined apathy as a neuropsychiatric syndrome characterised by a persistent lack of motivation that cannot be explained by diminished consciousness, cognitive deficits, or emotional distress. It is considered to be a multidimensional construct involving reduced motivation, goal-directed behaviour, and emotional indifference (Marin, 1996). These characteristics make apathy a critical focus in clinical assessment and intervention, given its substantial impact on daily functioning and overall quality of life.

Apathy has been consistently linked to significant impairments in psychosocial functioning. Individuals with apathy face difficulties in activities of daily living (Green et al., 2022; Tierney et al., 2018), reduced independence after hospital discharge (Arnould et al., 2015), and limited community integration (Cattelani et al., 2008). Apathy has also been associated with less progress in rehabilitation (Resnick et al., 1998), increased reliance on caregivers (Landes et al., 2001) and significant caregiver distress which can strain family dynamics (Marsh et al., 1998). Moreover, apathy has been associated with passive coping strategies (Finset & Andersson, 2000), poorer employment outcomes such as reduced working hours and financial independence(Bull et al., 2016; Funayama et al., 2022). These findings highlight the pervasive and multifaceted impact of apathy, reinforcing its importance as a focus for clinical assessment and intervention.

Apathy is prevalent across various other neurological conditions. A meta-analysis found that apathy affects 33% of individuals post-stroke (Zhang et al., 2023), and that the prevalence is approximately three times higher than depression (Caeiro et al., 2013). Similarly, 39.8% of individuals with Parkinson’s disease experience apathy (den Brok et al., 2015), and it was reported as the most common neuropsychiatric symptom in Alzheimer’s disease (Mega et al., 1996; Nobis & Husain, 2018; Zhao et al., 2016). However, our understanding of the prevalence of apathy following TBI is limited.

A recent review by Quang et al. (2024) explored factors associated with apathy in moderate-to-severe TBI and reported the limited role of injury severity and demographics. Instead, they highlighted the influence of factors like caregiver burden and self-efficacy. While this review emphasised the need for a multifaceted biopsychosocial approach to understanding apathy, it did not meta-analyse apathy prevalence, leaving the overall rate across studies unclear. Additionally, their focus on moderate-to-severe TBI limited insight into how apathy and its moderators vary across the full spectrum of injury severity.

Thus, despite recognition of its seriousness, the prevalence of apathy after TBI remains unclear and a robust meta-analysis addressing this is needed. Estimates of the prevalence of apathy in TBI vary widely, ranging from 16% (Zomeren & Burg, 1985) to 71% (Kant et al., 1998), making it difficult to estimate the scale of the problem and plan services accordingly. While the extent of this variability also hints at the existence of factors moderating the relationship between TBI and apathy, whether this is the case and which factors may be relevant remains unclear. This systematic review and meta-analysis aimed to address these important issues by providing a comprehensive estimate of apathy prevalence across mild, moderate, and severe TBI populations while identifying potential moderators influencing variability in reported rates.

## Methods

### Search Strategy

The protocol was pre-registered on PROSPERO (CRD42024552306). We performed comprehensive searches on the Ovid platform across three databases: APA PsycINFO, MEDLINE, and EMBASE. The search was conducted in August 2024 with no date restrictions. Exact search terms are provided in the supplementary appendix A. The search strategy included terms such as ‘traumatic brain injury’, ‘head injury’, ‘apathy’, ‘amotivation’ and ‘disinterest’. A wide-ranging selection of apathy-related terms was used (e.g. ‘loss of motivation, ‘indifference’).

### Eligibility Criteria and Screening

Eligibility criteria were defined using the Population, Intervention, Comparison, Outcomes, and Study (PICOS) framework:

- Population: Adults (≥18 years) with a history of TBI, assessed at least once for apathy.
- Intervention: Apathy assessed through clinical evaluation, structured interviews, or validated questionnaires.
- Comparison: Studies with or without comparison groups were eligible.
- Outcome: Prevalence of apathy reported as cases per sample.
- Study Design: Peer-reviewed primary research articles.

We included studies published in English only due to resource limitations but imposed no restrictions on publication date, geographic location, or care setting. We did not include studies which did not differentiate between TBI and other neurological conditions or acquired brain injury.

Search results were imported into Rayyan, an online screening platform. Two authors (JL and LHS) independently screened titles and abstracts for eligibility. Studies passing this initial screening underwent full-text review for final inclusion. Any disagreements were resolved through discussion or consultation with a third author (VB) when necessary.

### Data Extraction

Data were extracted independently by two authors (JL and LHS), with discrepancies resolved through discussion and consensus. A third author (VB) was consulted if necessary. Data were extracted into a pre-designed spreadsheet, capturing the following:

1. Study characteristics: Authors, publication year, study design, population and setting.
2. Sample demographics: Age, sex distribution, percentage of non-white participants and sample size.
3. Clinical characteristics: TBI severity, time since injury, and cause of TBI.
4. Apathy assessment: Measurement tool and rater (e.g., clinician, self-report, caregiver/informant).
5. Outcome: Prevalence of apathy (number of cases and sample size).

In cases where multiple apathy prevalence estimates were reported within a study, reviewers (JL and VB) selected the most appropriate frequency on a case-by-case basis. For instance, when both self-report and caregiver-report measures were available, the caregiver rating was chosen as this was the most frequently used method across studies. Additionally, when apathy prevalence was recorded at multiple time points, the mid-point estimate was used to ensure consistency.

### Quality Assessment

Risk of bias was evaluated using the JBI Critical Appraisal Checklist for Prevalence Studies which assesses various aspects of study validity such as measurement reliability, sample size and sampling methods (see supplementary appendix 2 for criteria). Assessments were conducted independently by two blinded authors (JL and LHS), with disagreements resolved through consensus.

The average score for study quality across all 18 studies was 8.6/10. 10 studies scored full marks for study quality and one scored below five. Full details of the quality assessment scores for each study can be found in supplementary appendix C.

### Statistical analysis

The meta-analysis was carried out using R (version 4.4.2) with the meta and metafor packages. Random-effects models were used to calculate the pooled prevalence of apathy after TBI. Heterogeneity was assessed using I² statistics to quantify the proportion of variability due to heterogeneity rather than sampling error. Heterogeneity is typical among prevalence studies and therefore a random effects model accounts for I^2^ ≥ 50%. Prevalence rates were transformed using a Freeman-Tukey double arcsine transformation to stabilize variance.

To assess potential publication bias, we constructed funnel plots and performed Egger’s regression test. Sensitivity analyses were completed to include a leave-one-out procedure, wherein each study was sequentially removed to examine its impact on the overall effect size.

Subgroup analyses were conducted to explore variations in prevalence based on study design, study location, setting, apathy measure and apathy rater. Meta-regressions were used to assess whether age, gender, study quality, TBI severity, length of time since TBI and cause of injury moderated the prevalence of apathy. All data and analysis code to reproduce the results in this study are freely available on the online archive at the following URL: https://github.com/lynchjess/Apathy-TBI-meta

## Results

### Search results

Out of 710 studies screened, 18 met the final inclusion criteria. The full screening and study selection process is detailed in the PRISMA flowchart (Figure 1). There was substantial agreement between both reviewers for the title and abstract screening (Cohen’s kappa = 0.72).

**Figure 1.**
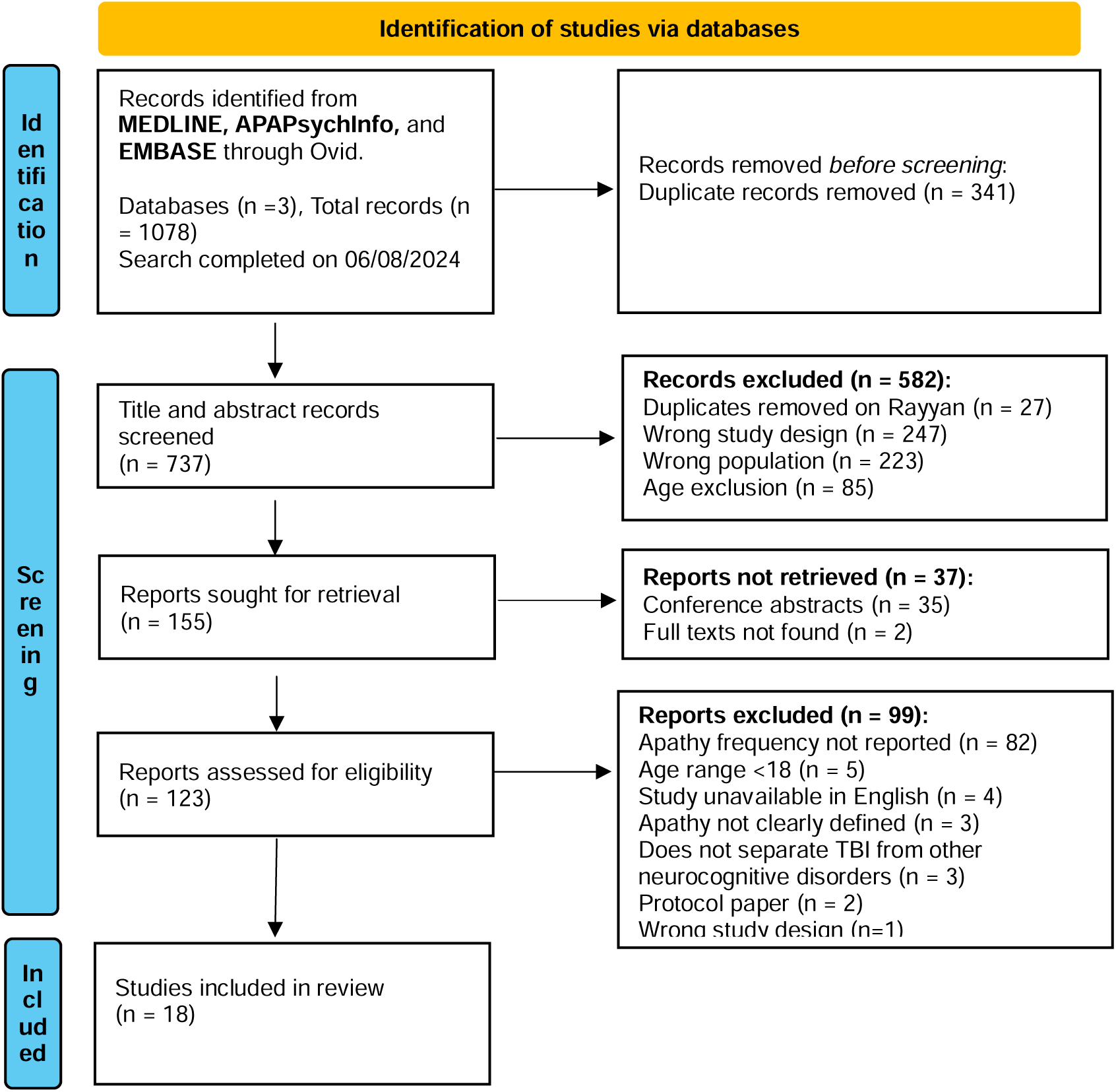
PRISMA flowchart for identification of included studies.

### Study characteristics

18 studies were included in the analysis, spanning publication years from 1991 to 2024, Table 1 outlines basic characteristics of each included study. The studies were conducted across five continents, with the majority based in North America (k = 6), Asia (k = 5) and Europe (k=5). Only two of the studies (both in North America) reported details on proportion of non-white participants. A total of 1,136 participants with a TBI were included, with a mean of 63 participants per study. The mean age of participants was 37.8 years, and males comprised 79% of the sample, reflecting the well-documented gender imbalance in reported TBI rates (Bruns Jr. & Hauser, 2003).

**Table 1.**
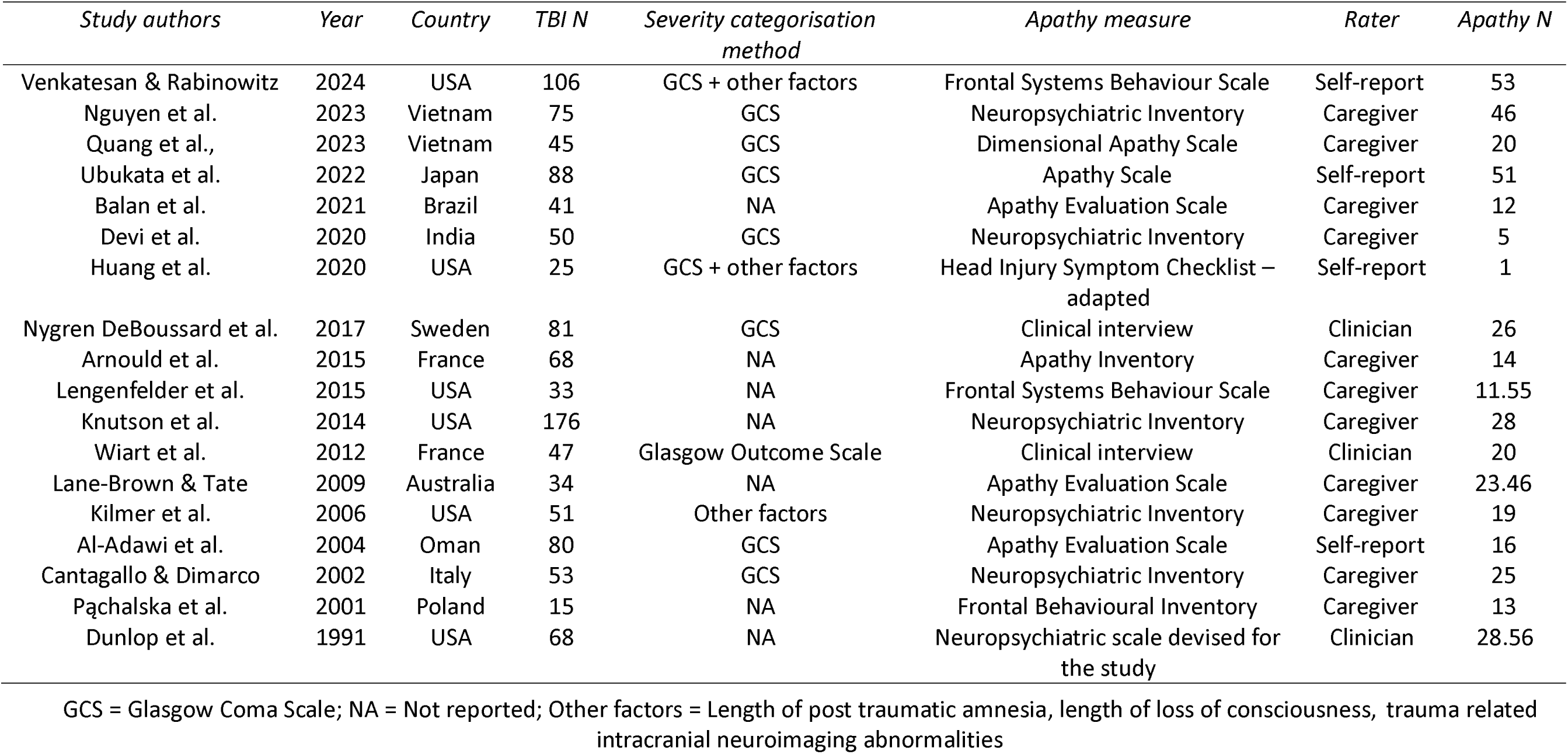
Characteristics of 18 included studies.

One study (Nygren DeBoussard et al., 2017) provided demographic data for a larger sample (n=114) than the 81 that were assessed for apathy, limiting the demographic details we could report for this study. 16 studies reported the TBI severity of their sample, which overall was found to be 10.9% mild, 31.5% moderate - severe and 50.7% severe. Seven of the studies used the Glasgow Coma Scale (GCS) to categorise severity: mild (score 13-15), moderate (9- 12), and severe (3-8). Two studies used the GCS alongside other factors, such as the duration of loss of consciousness, post-traumatic amnesia, and intracranial neuroimaging abnormalities. One study combined these factors without relying on the GCS, while another used the Glasgow Outcome Scale. Seven studies did not specify their method of categorisation.

Eleven studies reported TBI causes: transport accidents (74.5%), falls (11.36%), combat (4.3%), assaults (4.5%), and other causes (3.8%). Seventeen studies reported a mean time since TBI of 43.4 months. Fourteen studies did not specify whether participants had a single or multiple TBIs. Of the four that did, two included only single TBI cases, while the other two included participants with varying numbers of TBIs.

Apathy was assessed using a variety of measures, including the Neuropsychiatric Inventory (k = 5), Apathy Evaluation Scale (k = 3), Frontal Symptoms Behaviours Scale (k = 2), Dimensional Apathy Scale (k = 1), Apathy Scale (k=1), Head Injury Symptom Checklist (k=1), Frontal Behavioural Inventory (k=1) and Apathy Inventory (k=1). 15 of these were validated measures and the remaining three were either assessed via clinical evaluation or a scale specifically devised for the study. Most measures were caregiver-rated (k = 11), while four were self-reported and three were rated by clinicians. Reported apathy prevalence rates ranged widely, from 4% to 87%.

### Main Findings

The pooled prevalence of apathy after TBI was 37.58% (95% CI: 28.45–47.15%) under a random-effects model (Figure 2). Heterogeneity was high (I² = 90.3%, 95% CI: 86.2–93.2%), with a significant test of heterogeneity (Q = 174.88, p < 0.001) and between-study variance (τ² = 0.037, 95% CI: 0.021–0.098). Study weights ranged from 4.4% to 6.1%, indicating no single study disproportionately influenced the estimate. A Freeman-Tukey double arcsine transformation and Clopper-Pearson method were applied to address extreme values and calculate confidence intervals.

**Figure 2.**
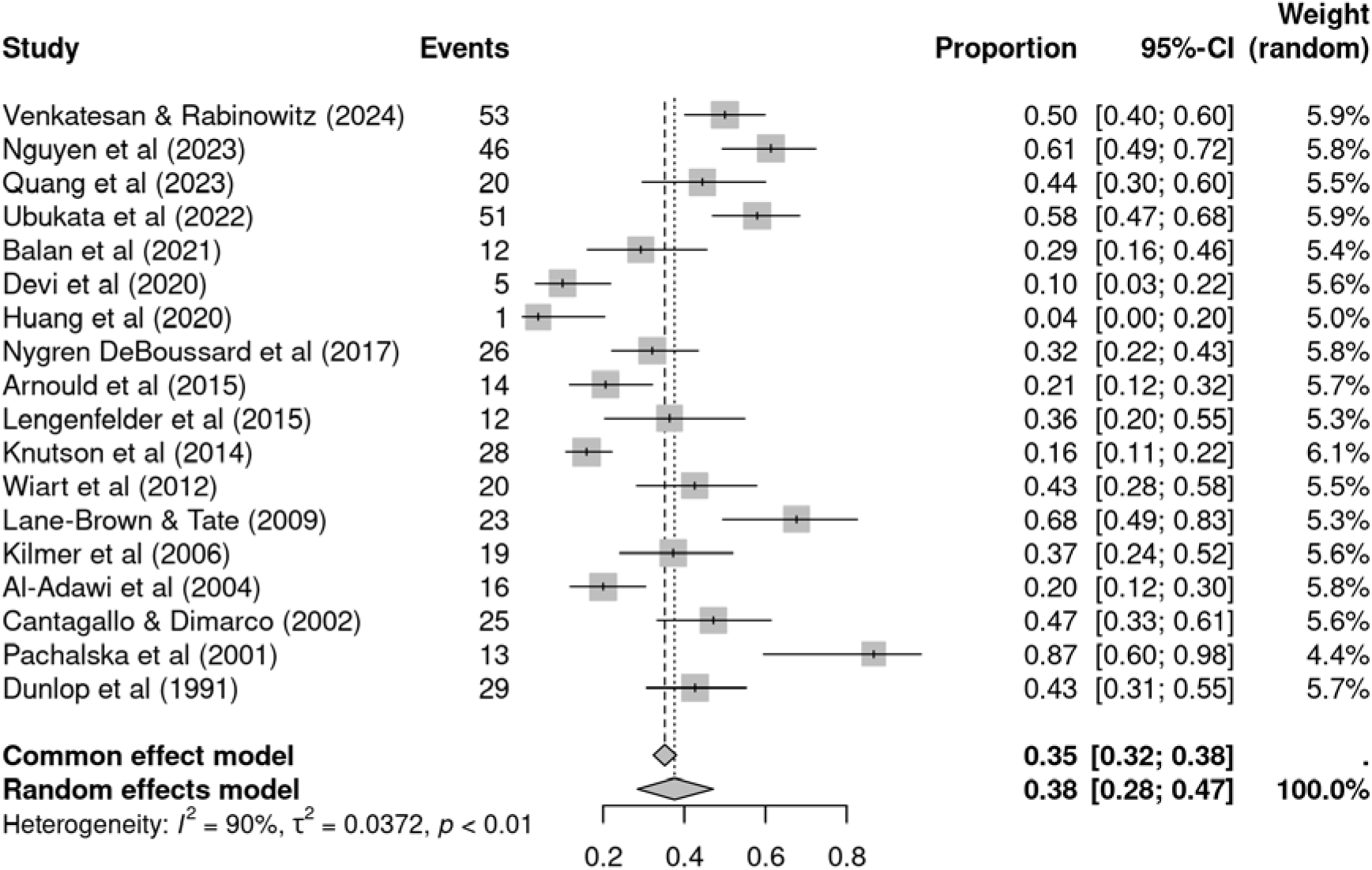
Forest plot of overall apathy prevalence across 18 included studies.

### Subgroup analyses

We conducted subgroup analyses for study design, setting, population, continent, apathy measure, validated measures and apathy rater; see Table 2 for full results. Groups with single studies only were excluded. The only significant difference was found was in population subgroups, where veterans had a lower apathy prevalence than the general public (p <0.001). No other subgroups were found to significantly moderate the apathy prevalence.

**Table 2.** Subgroup Analyses of Apathy Prevalence in TBI Populations.

| *Subgroup* | *Category (k)* | *Prevalence (%)* | *95% CI* | *p* |
| --- | --- | --- | --- | --- |
| Study Design | Cross-sectional (14) | 40.09 | 28.97–51.72 | 0.306 |
|  | Other designs (4) | 29.66 | 15.20–46.46 |  |
| Setting | Outpatient (8) | 43.23 | 29.24–57.78 | 0.600 |
|  | Inpatient (5) | 40.45 | 24.30–57.69 |  |
|  | Study registry database (3) | 30.69 | 13.46–51.21 |  |
| Population | General public (16) | 41.52 | 32.77–50.54 | < 0.001 |
|  | Veterans (2) | 10.92 | 2.47–23.56 |  |
| Continent | Europe (5) | 43.18 | 26.89–60.22 | 0.513 |
|  | Asia (5) | 37.46 | 17.94–59.30 |  |
|  | North America (6) | 29.87 | 15.95–45.90 |  |
| Apathy Measure | Frontal Systems Behaviour Scale (2) | 45.12 | 32.63–57.92 | 0.672 |
|  | Apathy Evaluation Scale (3) | 37.66 | 13.20–65.91 |  |
|  | Clinical Interview (2) | 36.23 | 26.47–46.58 |  |
|  | Neuropsychiatric Inventory (5) | 32.71 | 14.47–54.09 |  |
| Validated Measures | Validated (15) | 37.37 | 26.46–48.94 | 0.905 |
|  | Non-validated (3) | 38.27 | 31.12–45.67 |  |
| Apathy Rater | Caregiver (11) | 39.60 | 26.45–53.52 | 0.836 |
|  | Self-report (4) | 31.49 | 11.61–55.60 |  |
|  | Clinician (3) | 38.27 | 31.12–45.67 |  |

### Meta regression analyses

We also ran several meta-regressions to explore the impact of further demographic factors (age and sex), TBI factors (severity, time since TBI and cause of TBI) and study factors (year published and study quality) on apathy prevalence.

#### Demographic factors

A mixed-effects meta-regression model found no significant relationship between mean age and apathy prevalence (β = 0.001, p = 0.930). An analysis of 17 studies revealed a significant effect of sex, with males showing a lower prevalence of apathy (β = −0.91, p = 0.03). The model explained 10.56% of the heterogeneity, but residual heterogeneity remained high (I² = 90.03%).

#### TBI factors - severity

Three meta-regression analyses were conducted to examine whether TBI severity moderated the relationship between TBI and apathy prevalence. 15 studies were included, with three excluded due to missing data.

A significant negative association was found between mild TBI and apathy prevalence (β = - 0.40, p = 0.044), suggesting that individuals with mild TBI experience lower apathy prevalence compared to other severity levels. The model explained a modest proportion of variability (R² = 5.16%), though residual heterogeneity remained high (I² = 87.79%). No significant associations were found for moderate-severe TBI (β = 0.10, p = 0.469) or severe TBI (β = 0.09, p = 0.473) with apathy prevalence.

#### TBI factors - time since TBI and cause of TBI

A meta-regression analysis investigating the relationship between the mean months since TBI and apathy prevalence revealed no significant association (β = 0.002, p = 0.130). Regarding the cause of TBI, additional meta-regression analyses were conducted to assess how specific causes influenced apathy prevalence. For transport accidents, an analysis of 11 studies demonstrated a significant positive association, indicating that individuals with TBI resulting from transport accidents exhibited higher apathy prevalence (β = 0.417, p = 0.027). This model explained 26.02% of the variability and residual heterogeneity remained high (I² = 89.56).

When exploring the same 11 studies, no significant relationship was identified between falls as a cause of TBI and apathy prevalence (β = 0.125, p = 0.908). Similarly, an analysis focusing on assault as a cause of TBI, which included nine studies where this was reported, found no significant association with apathy prevalence (β = 1.150, p = 0.397).

#### Study factors

A meta-regression analysis found no significant relationship between the year of study publication and apathy prevalence (β = −0.004, p = 0.428). Similarly, no significant effect was found for study quality (β = −0.014, p = 0.601).

### Publication bias and sensitivity analysis

The Egger’s test for funnel plot asymmetry suggests no statistically significant evidence of publication bias in the meta-analysis (t = 1.13, df = 16, p = 0.274), see Figure 3 for funnel plot. The bias estimate is 3.250 (SE = 2.871), indicating a slight tendency for asymmetry, but no significant asymmetry or small-study effects.

**Figure 3.**
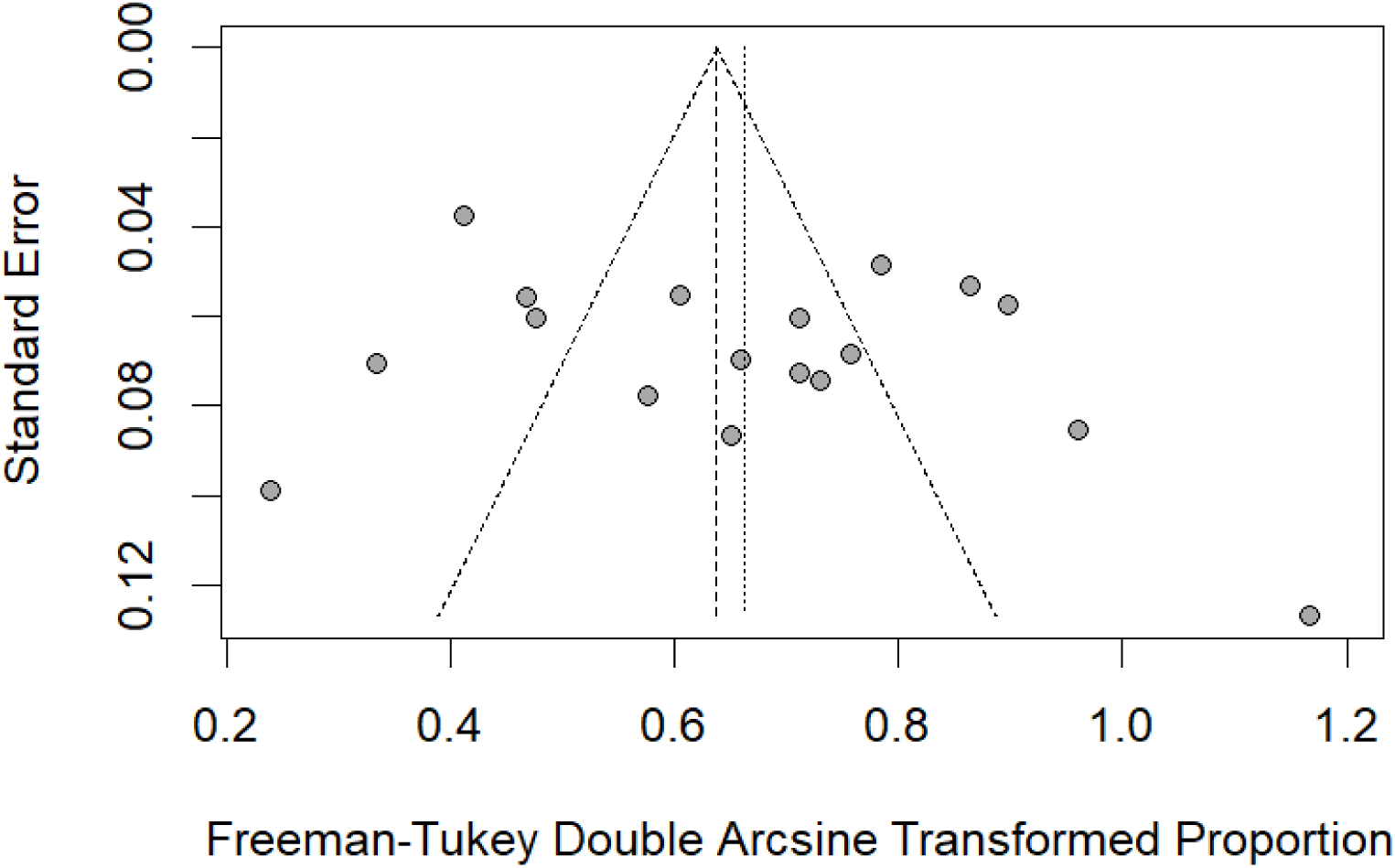
Funnel plot.

#### Outlier and influence analysis

Outlier analysis identified seven studies as influential and removed them from the meta-analysis, including Nguyen et al., (2023), Devi et al., (2020) and Knutson et al., (2014). After excluding these outliers, the random-effects model was based on 11 studies with 673 observations and 246 events. The pooled estimate for apathy prevalence was 36.11% (95% CI: 29.37–43.13), with substantial heterogeneity (I² = 70.4%, τ² = 0.009). The heterogeneity test was significant (Q = 33.77, p < 0.001).

Sensitivity analysis using leave-one-out diagnostics showed minor fluctuations in the pooled prevalence (ranging from 35.34% to 39.48%), but these did not affect statistical significance or overall heterogeneity (I² = 90.3%), indicating the robustness of the meta-analytic results.

The influence diagnostics suggest that some studies, such as Knutson et al., (2014) and Nguyen et al., (2023) have significant effects on the pooled effect size and heterogeneity. Baujat diagnostics further identified Knutson et al., (2014) as a major contributor to heterogeneity. However, despite these variations, the overall conclusion about the prevalence of apathy remains robust across different models and analyses. An influence chart is available in the supplementary materials.

## Discussion

To our knowledge, this is the first review to meta-analyse the pooled prevalence of apathy in individuals with a history of TBI. We also examined potential moderators, including demographic characteristics, TBI-related factors, and methods of apathy assessment. Drawing on data from 18 studies published between 1991 and 2024, we found an overall prevalence of 37.6%, emphasising the high occurrence of apathy in this population. The prevalence estimate remained robust with little change in the overall estimate in subsequent sensitivity analyses.

This prevalence rate is comparable to other neurological conditions, such as 33% in stroke (Zhang et al., 2023) and 39.8% in Parkinson’s disease (den Brok et al., 2015), although it is lower than the 49% prevalence found in Alzheimer’s disease (Zhao et al., 2016). These findings indicate that over a third of individuals with TBI are likely to experience apathy. Given its significant negative impact on psychosocial functioning, rehabilitation outcomes, and family well-being, this underlines the need for greater clinical attention to apathy as a critical issue in TBI recovery.

Significant heterogeneity was observed across studies, with apathy frequency ranging from 4% to 87%. This wide variability suggests the influence of various moderating factors. Our subgroup analyses and meta-regressions identified four factors that significantly influenced apathy prevalence: cause of injury, TBI severity, sex and population type. Specifically, transport accidents were associated with higher apathy prevalence, while mild TBI, male sex, and veteran status were associated with lower apathy prevalence. We will consider each of these in turn.

We found that males had a significantly lower prevalence of apathy compared to females. Previous studies, including a large cohort study and meta-analysis, have found that females reported a higher symptom burden and more mental health difficulties following TBI (Farace & Alves, 2000; Mikolić et al., 2021). This difference was more pronounced after mild TBI than after severe TBI (Mikolić et al., 2021; Starkey et al., 2022). The mechanisms underlying these disparities in sex remain unclear, with some clinical opinions contrasting this and suggesting that females tend to experience better outcomes after TBI. While biological factors may contribute to these differences, an additional explanation could be that females are more likely to self-report symptoms or have greater self-awareness of TBI-related deficits (Barsky et al., 2001; Niemeier et al., 2014). Interestingly, a review on apathy after TBI (Quang et al., 2024) found no significant sex differences. However, this review focused only on moderate-to-severe TBI and may not reflect differences that emerge more clearly in mild TBI, which our study includes. Given the male predominance in TBI cases (79% in our sample), further research is needed to explore sex-specific differences in apathy and their clinical implications. No significant impact of other demographic factors, such as age, was found on apathy prevalence in this review.

Regarding TBI severity, we conducted three meta-regressions to examine whether the severity of TBI (mild, moderate-severe, and severe) moderated the relationship between TBI and apathy prevalence. Our results indicated that only mild TBI had a significant moderating effect, suggesting that individuals with mild TBI may be less likely to experience apathy. However, this effect was small, and substantial heterogeneity remained, indicating that additional factors may contribute to apathy in this group. The lack of significant findings for moderate-to-severe and severe TBI suggests that apathy prevalence may not increase in direct proportion to injury severity alone. Neuropsychiatric outcomes after mild TBI are influenced by a complex, multifactorial aetiology, with psychological mechanisms such as coping styles or pre-injury mental health difficulties potentially playing a significant role in this group (Mooney et al., 2005; Ponsford et al., 2000; van der Horn et al., 2020). Given that mild TBI accounted for only 10% of the participants in this review, these findings should be interpreted with caution and further research on apathy in mild TBI is needed.

Road traffic accidents were found to moderate the relationship between TBI and apathy, suggesting that individuals with traffic accident-related TBIs may be more likely to experience apathy. One potential explanation for this finding is that road traffic accidents may lead to more complex neurological damage or additional injury-related factors that influence apathy. In high-income countries, road traffic accidents account for the highest proportion of TBI-related hospitalisations (Hyder et al., 2007), indicating that these injuries often necessitate more intensive medical intervention. Furthermore, TBIs resulting from road traffic accidents tend to be more severe than those caused by other incidents (Rahman et al., 2025). The increased severity of injury and the heightened burden of hospitalisation following road traffic accidents may contribute to the higher prevalence of apathy observed in this group.

Veterans had a significantly lower prevalence of apathy (10.9%) compared to the general population (41.5%). However, this result should be interpreted cautiously, given the small number of studies on veterans (*k* = 2). Notably, both veteran cohorts were composed entirely of males. Given that we found males had a significantly lower prevalence of apathy than females, one possibility is that this difference is explained entirely by sex. Alternatively, the lower prevalence in veterans could be attributed to other factors, such as distinct TBI aetiologies, the effects of military training or rehabilitation, or differences in symptom reporting. One of these studies used self-report measures, while the other relied on caregiver ratings. Some research suggests that veterans may underreport psychological symptoms due to cultural influences such as military norms around emotional resilience and concerns about stigma (Hoge et al., 2004; Vogt et al., 2014). If underreporting is a factor, the true prevalence of apathy in veterans may be underestimated. Further research using clinician-rated measures may help clarify whether the lower prevalence reflects a genuine difference or is influenced by reporting biases. More research with larger veteran cohorts, particularly those including females, is necessary to confirm and expand upon these findings.

We found no evidence that the length of time since injury influenced apathy prevalence, suggesting that apathy may persist over time. This might indicate that apathy is a chronic symptom, possibly driven by neurobiological changes affecting brain regions involved in motivation and emotion. The lack of significant change over time may also reflect insufficient clinical focus on apathy post-TBI. Notably, a systematic review on interventions for post-TBI apathy identified only one randomised controlled trial addressing this symptom (Lane□Brown & Tate, 2009). This study examined cranial electrotherapy stimulation but lacked between-group analyses, preventing conclusions on effectiveness. This highlights the scarcity of evidence on effective treatments.

Our analysis found that apathy prevalence remained consistent across different study designs, settings, and continents. Likewise, prevalence estimates were not influenced by the type of apathy measure used or whether it was validated. This consistency suggests that our prevalence estimate is robust, regardless of assessment approach.

The study has several methodological strengths. By pooling data from multiple studies and using meta-analytic techniques, we achieved high statistical power, which allowed for more precise and reliable estimates of apathy prevalence. The use of subgroup analyses and meta-regressions further enabled us to explore potential moderating factors, helping to address the considerable variability in reported apathy rates. Moreover, the inclusion of studies spanning different TBI severities strengthens the generalisability of our findings across a wide range of patient populations.

Despite these strengths, several limitations warrant consideration. While the review included studies from five continents, no studies from Africa and only one from Oceania were identified, narrowing the global applicability of the findings. We also only included studies published in English. Moreover, most studies failed to report the ethnicity of their samples, limiting exploration of the influence of non-white representation. The findings are further constrained by the underrepresentation of certain groups, such as females and individuals with mild TBI, hindering the understanding of sex-specific differences and limiting the applicability to milder forms of TBI. Furthermore, the categorisation of TBI severity was not consistently reported across studies, and variability in classification systems could have impacted the results. Lastly, while our meta-analysis identified several factors that are likely to explain some of the variability in prevalence estimates, it is notable that these estimates were still extremely wide (ranging from 4% to 87%). It may be that other factors not identified as part of this analysis are contributing to this variability.

Our findings highlight the high prevalence of apathy following TBI, yet treatment options remain limited. In dementia, acetylcholinesterase inhibitors have shown the strongest evidence for reducing apathy (Berman et al., 2012), while motivation-based behaviour therapy has demonstrated benefits in a single-case experimental study (Lane-Brown & Tate, 2010). Multi-sensory stimulation and music therapy have also shown promise in dementia populations (Holmes et al., 2006; Verkaik et al., 2005). Given the multifaceted nature of apathy, clinical research should prioritise developing and evaluating pharmacological, psychological, and rehabilitative interventions tailored to TBI-related apathy.

In conclusion, this meta-analysis is the first to systematically quantify the prevalence of apathy in individuals with TBI and identify the influence of moderating factors. Our findings demonstrate that apathy is prevalent following TBI, affecting over one-third of individuals in the reviewed studies. It has highlighted significant variability in prevalence rates, whilst also identifying several factors that may contribute to this variation. Specifically, transport accidents were associated with higher apathy prevalence, while mild TBI, male sex, and veteran status were associated with lower apathy prevalence. These findings underscore the importance of recognising apathy as a key aspect of TBI recovery and emphasise the need for greater clinical attention to its assessment and management.

## Data Availability

All data produced are available online at in the study github archive

https://github.com/lynchjess/Apathy-TBI-meta

