## Supplementary Material for "Prevalence and moderators of apathy after traumatic brain injury: a systematic review and meta-analysis"

**Appendix A:** Search terms

"apathy" OR "abulia" OR "amotivation" OR "avolition" OR "indifference" OR "emotional indifferen*" OR "disinterest" OR "neuropsychiatric inventory" OR "frontal lobe personality scale" OR "Lille apathy rating scale" OR "frontal system behavior scale" OR "frontal system behaviour scale" OR "key behavior change inventory" OR "key behaviour change inventory" OR "apathy evaluation scale" OR "apathy scale" OR "irritability-apathy scale" OR "irritability apathy scale" OR "dimensional apathy scale" OR "apathy motivation index" OR "reduced interest" OR "reduced drive" OR "reduced volition" OR "reduced motivation" OR "diminished interest" OR "diminished drive" OR "diminished volition" OR "diminished motivation" OR "loss of interest" OR "loss of drive" OR "loss of volition" OR "loss of motivation"

*AND*

"traumatic brain injur*" OR TBI OR concussion OR "craniocerebral trauma" OR "head injury" OR "Craniocerebral Trauma" [Mesh]

**Appendix B:** JBI Critical Appraisal Checklist for studies reporting prevalence data

|  | Yes | No | Unclear | Not applicable |
| --- | --- | --- | --- | --- |
| 1. Was the sample frame appropriate to address the target population? | □ | □ | □ | □ |
| 2. Were study participants sampled in an appropriate way? | □ | □ | □ | □ |
| 3. Was the sample size adequate? | □ | □ | □ | □ |
| 4. Were the study subjects and the setting described in detail? | □ | □ | □ | □ |
| 5. Was the data analysis conducted with sufficient coverage of the identified sample? | □ | □ | □ | □ |
| 6. Were valid methods used for the identification of the condition? | □ | □ | □ | □ |
| 7. Was the condition measured in a standard, reliable way for all participants? | □ | □ | □ | □ |
| 8. Was there appropriate statistical analysis? | □ | □ | □ | □ |
| 9. Are all important confounding factors and subgroups differences identified and accounted for? | □ | □ | □ | □ |
| 10. Were subpopulations identified using objective criteria? | □ | □ | □ | □ |

**Appendix C:** Study quality assessment

| **Year** | **Study** | **Q1** | **Q2** | **Q3** | **Q4** | **Q5** | **Q6** | **Q7** | **Q8** | **Q9** | **Q10** | **Summary score** |
| --- | --- | --- | --- | --- | --- | --- | --- | --- | --- | --- | --- | --- |
| 2024 | Venkatesan & Rabinowitz | 1 | 1 | 1 | 1 | 1 | 1 | 1 | 1 | 1 | 1 | **10** |
| 2023 | Nguyen et al. | 1 | 1 | 1 | 1 | 1 | 1 | 1 | 1 | 1 | 1 | **10** |
| 2023 | Quang et al. | 1 | 1 | 1 | 1 | 1 | 1 | 1 | 1 | 1 | 1 | **10** |
| 2022 | Ubukata et al. | 1 | 1 | 1 | 1 | 1 | 1 | 1 | 1 | 1 | 1 | **10** |
| 2021 | Balan et al. | 1 | 1 | 1 | 0 | 1 | 1 | 1 | 1 | 0 | 1 | **8** |
| 2020 | Devi et al. | 1 | 1 | 1 | 1 | 1 | 1 | 1 | 1 | 1 | 1 | **10** |
| 2020 | Huang et al. | 1 | 1 | 1 | 1 | 1 | 1 | 1 | 1 | 1 | 1 | **10** |
| 2017 | Nygren DeBoussard et al. | 1 | 1 | 1 | 1 | 0 | 0 | 00 | 1 | 1 | 00 | **6** |
| 2015 | Arnould et al. | 1 | 1 | 1 | 1 | 1 | 1 | 1 | 1 | 1 | 1 | **10** |
| 2015 | Lengenfelder et al. | 1 | 00 | 1 | 0 | 1 | 1 | 1 | 1 | 0 | 1 | **7** |
| 2014 | Knutson et al. | 1 | 1 | 1 | 0 | 1 | 1 | 1 | 1 | 0 | 000 | **7** |
| 2012 | Wiart et al. | 1 | 1 | 1 | 1 | 1 | 0 | 0 | 1 | 1 | 1 | **8** |
| 2009 | Lane-Brown & Tate | 1 | 1 | 1 | 1 | 1 | 1 | 1 | 1 | 1 | 1 | **10** |
| 2006 | Kilmer et al. | 1 | 1 | 1 | 1 | 1 | 1 | 1 | 1 | 1 | 1 | **10** |
| 2004 | Al-Adawi et al. | 1 | 1 | 1 | 1 | 1 | 1 | 1 | 1 | 1 | 1 | **10** |
| 2002 | Cantagallo & Dimarco | 1 | 1 | 1 | 1 | 1 | 1 | 1 | 1 | 0 | 1 | **9** |
| 2001 | Pachalska et al. | 1 | 1 | 0 | 0 | 1 | 1 | 1 | 1 | 0 | 000 | **6** |
| 1991 | Dunlop et al. | 1 | 0 | 1 | 0 | 1 | 00 | 0 | 1 | 00 | 00 | **4** |

Notes: 1 = Yes, 0 = No, 00 = Unclear, 000 = NA.

**Appendix D:** Influence analysis

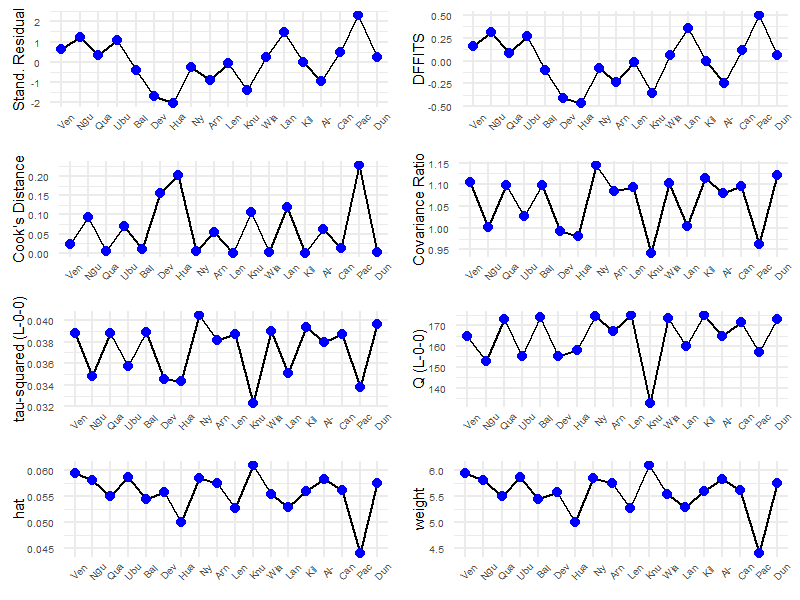
